# Targeting TGF-β pathway with COVID-19 Drug Candidate ARTIVeda/PulmoHeal Accelerates Recovery from Mild-Moderate COVID-19

**DOI:** 10.1101/2021.01.24.21250418

**Authors:** Vuong Trieu, Saran Saund, Prashant V. Rahate, Viljay B. Barge, K. Sunil Nalk, Hitesh Windlass, Fatih M. Uckun

**Affiliations:** Oncotelic Inc., Agoura Hills, CA 91301, USA; Seven Star Hospital, Nagpur-440009, Maharashtra, India; Government Medical College and Chhatrapati Pramila Raje Hospital, Chowk, Kolhapur- 431001, Maharashtra, India; Government Medical College and Government General Hospital, Srikakulam-532001, Andhra Pradesh, India; Windlas Biotech Pvt. Ltd., Dehradun UR 248110, India; Ares Pharmaceuticals, LLC, St. Paul, MN 55110, USA

## Abstract

Our COVID-19 drug candidate ARTIVeda™/PulmoHeal is a novel gelatin capsule formulation of the Artemisia extract Ayurveda for oral delivery of TGF-β targeting anti-malaria phytomedicine Artemisinin with documented anti-inflammatory and anti-SARS-CoV-2 activity. Here we report the safety and efficacy of ARTIVeda™ in adult COVID-19 patients with symptomatic mild-moderate COVID-19, who were treated in a randomized, open-label Phase IV study in Bangalore, Karnataka, India (Clinical Trials Registry India identifier: CTRI/2020/09/028044). ARTIVeda showed a very favorable safety profile, and the only ARTIVeda-related adverse events were transient mild rash and mild hypertension. Notably, ARTIVeda, when added to the SOC, accelerated the recovery of patients with mild-moderate COVID-19. While all patients were symptomatic at baseline (WHO score = 2-4), 31 of 39 (79.5%) of patients treated with ARTIVeda plus SOC became asymptomatic (WHO score = 1) by the end of the 5-day therapy, including 10 of 10 patients with severe dry cough 7 of 7 patients with severe fever. By comparison, 12 of 21 control patients (57.1%) treated with SOC alone became asymptomatic on day 5 (P=0.028, Fisher’s exact test). This clinical benefit was particularly evident when the treatment outcomes of hospitalized COVID-19 patients (WHO score = 4) treated with SOC alone versus SOC plus ARTIVeda were compared. The median time to becoming asymptomatic was only 5 days for the SOC plus ARTIVeda group (N=18) but 14 days for the SOC alone group (N=10) (P=0.004, Log-rank test). These data provide clinical proof of concept that targeting the TGF-β pathway with ARTIVeda may contribute to a faster recovery of patients with mild-moderate COVID-19 when administered early in the course of their disease.

## Introduction

Effective drugs are needed to prevent the potentially deadly complications of COVID-19 (1-3) and thereby reduce its fatality rate has become of the main causes of death (4-9). Artemisinin is an anti-inflammatory phytomedicine with broad-spectrum antiviral activity and a commonly used anti-malaria drug (10-12). Artemisinin and some of its chemical derivatives exhibit broad-spectrum antiviral activity against pathogenic human viruses (11,12). Notably, recent docking studies indicated that Artemisinin and its derivative Artesunate could bind the SARS-CoV-2 spike protein in a way that would interfere with its docking onto the human ACE2 receptor protein, which is the required first step in the host infection process of the coronavirus disease 2019 (COVID-19) (13,14). Studies by Cao et al.(15) and Gilmore et al. confirmed the antiviral activity of Artemisinin and its derivatives against SARS-2-CoV-2 (16). Its documented anti-SARS-CoV-2 activity has been attributed to its ability to inhibit spike-protein mediated and TGF-β-dependent early steps in the infection process (12). It is noteworthy that the incidence of COVID-19 and related deaths have been very low in countries with regular use of artemisinin-based anti-malaria drugs (17). Consequently, Artemisinin has recently been repurposed as a potential COVID-19 drug (12). Li et al. reported the results from an open-label non-randomized study in which 41 COVID-19 patients received either standard of care (SOC) therapy (control) or SOC combined with Artemisinin plus piperaquine (AP) (18). Patients in the AP group showed a faster clearance of SARS-CoV-2 than control patients.

Our COVID-19 drug candidate ARTIVeda™ is a novel gelatin capsule formulation for oral delivery of Artemisinin. The purpose of the present study was to evaluate the safety and efficacy of ARTIVeda™ in adult COVID-19 patients with symptomatic mild-moderate COVID-19, who were treated in a randomized, open-label Phase IV study in Bangalore, Karnataka, India (Clinical Trials Registry India identifier: CTRI/2020/09/028044). ARTIVeda showed a very favorable safety profile and significantly accelerated the recovery of patients with mild-moderate COVID-19 when added to the SOC. For patients with moderate COVID-19 (WHO Score = 4), the median time to becoming asymptomatic was only 5 days for the SOC plus ARTIVeda group (N=18) but 14 days for the SOC alone group (N=10) (P=0.004, Log-rank test). These findings provide clinical proof of concept that the use of ARTIVeda as an adjunct to the SOC may result in faster recovery of patients with mild-moderate COVID-19.

## Materials and Methods

### ARTIVeda™/PulmoHeal, a novel gelatin capsule formulation of the Artemisia extract Ayurveda for oral delivery of Artemisinin

The product, ArtiVeda™/PulmoHeal (License # UK.AY-401/2018, Ministry of AYUSH, India), is a novel gelatin capsule formulation of the Artemisia extract Ayurveda for oral delivery of the active ingredient Artemisinin for treatment of COVID-19. This dual-function COVID-19 drug candidate has potent antiviral activity against SARS-CoV-2 and is expected to mitigate the TGF-β mediated inflammatory injury associated with the cytokine storm and viral sepsis in critically ill COVID-19 patients.

### ARTI-19 Clinical Study

The ARTI-19 is a randomized, open-label Phase IV study was designed to evaluate the safety and efficacy of ARTIVeda when used in combination with the standard of care (SOC) in mild-moderate COVID-19 patients in Bangalore, Karnataka, India. The official title of the study protocol is “A Prospective, Randomized, Multi-center, Open-label, Interventional Study to Evaluate the Safety and Efficacy of Artemisinin 500 mg capsule in Treatment of Adult Subjects with COVID-19”. The ARTI-19 trial was cleared by Indian regulatory authorities and is registered under the Clinical Trials Registry India (CTRI) with three active sites (CTRI/2020/09/028044). This study is sponsored by Oncotelic, Inc. (USA) and Windlas Biotech Private Limited (India). The execution of the trial has been contracted to the Clinical Research Organization (CRO) Abiogenesis Clinpharm Private Limited (Hyderabad, India). The primary endpoint of the study is time to recovery from the signs and symptoms of COVID-19.

The study will initially enroll 120 adult patients with RT-PCR confirmed and symptomatic mild-moderate COVID-19 not requiring any oxygen therapy (i.e., scores of 2-4 on the 10-point WHO Clinical Progression Scale) who will be randomized to either standard of care (SOC) plus ARTIVeda or SOC alone. Consenting registered patients are randomly divided by computer-generated randomization sequence 2:1 into the experimental arm (Group 1: ARTIVeda plus SOC): control arm (Group 2: SOC alone). This report details the clinical data in the first 60 randomized patients who were treated at the following 3 sites: 1) Site 203: Government Medical College & Government General Hospital, Srikakulam, ANDHRA PRADESH; 2) Site 202: Rajarshi Chhatrapati Shahu Maharaj Government Medical College and Chhatrapati Pramila Raje Hospital, MAHARASHTRA; and 3) Site 201: Seven Star Hospital, MAHARASHTRA. The SOC was generally as described in the national clinical management protocol of India: “COVID-19, Government of India Ministry of Health and Family Welfare Directorate General of Health Services (EMR Division)”. There were site-to-site variations in choice of SOC drugs. Site 201 employed Remdesivir, prednisolone, low molecular weight heparin, and doxycycline as the key components of the SOC. Azithromycin, dexamethasone were part of the SOC used at Site 202. Site 203 SOC included Ivermectin, doxycycline, and inhaled steroids.

For patient eligibility in the trial, the Inclusion Criteria are 1. Confirmed case of COVID-19 infection by laboratory tests for COVID-19; 2. Age limit: 21 to 60 years of age male, or non-pregnant or non-lactating female; 3. Patients with oxygen saturation higher than 95% and without any requirement of oxygen therapy or assisted ventilation; 4. Patients willing to give their informed consent to participate in the clinical trial. Patients are excluded if they have one or more of the following Exclusion Criteria: 1. COVID-19 positive patients above 60 years of age or below 21 years; 2. Patients on Immuno-suppression therapy; 3.Patients with associated renal or hepatic impairment; 4. Pregnant women or lactating mothers; 5.

Patients in an advanced stage of disease requiring emergency medical intervention like pneumonia, bronchial asthma, organ failure; 6. Patients whom ventilator support is required; 7. Patients not willing to give their informed consent to participate in the clinical trial; 8. Patients with the following co-morbidities: insulin-dependent diabetes, hypertension with cardiac symptoms, morbid obesity with diabetes and/or hypertension. Uncontrolled diabetes. Uncontrolled hypertension.

### Ethics Statement and Approval

ARTI-19 is being performed in adherence to the Good Clinical Practice (GCP) guidelines, Ministry of Ayurveda, Yoga and Naturopathy, Unani, Siddha and Homoeopathy (AYUSH) guidelines, and Indian Council of Medical Research. The study was approved by Ethics Committees at the three participating institutions: 1) Government Medical College & Government General Hospital, Srikakulam, ANDHRA PRADESH. 2) Rajarshi Chhatrapati Shahu Maharaj Government Medical College and Chhatrapati Pramila Raje Hospital, MAHARASHTRA. And 3) Seven Star Hospital, MAHARASHTRA.

## Results

### Patient characteristics

We present data from 60 Asian patients with COVID-19 who were treated on the ART-19 study between October 8, 2020 and November 21, 2020. The date for data cut off was November 5, 2020. 39 patients were randomized to the experimental arm and received ARTIVeda as an adjunct to SOC, whereas 21 patients were randomized to the control arm and received SOC without ARTIVeda. The baseline patient characteristics are shown in **Table 1**. The median age was 44.5 years (Range: 18-59 years). 38 patients (63.3%) were male and 22 patients (36.7%) were female. All 60 patients had RT-PCR confirmed symptomatic mild-moderate COVID-19 with a score of 2-4 on the 10-point WHO clinical progression scale not requiring any oxygen therapy, of whom 28 (46.7%) were hospitalized patients with a score of 4 (**Table 1**). There were no significant differences in the baseline characteristics of patients on the experimental versus control arms of the protocol.

**Table 1.**
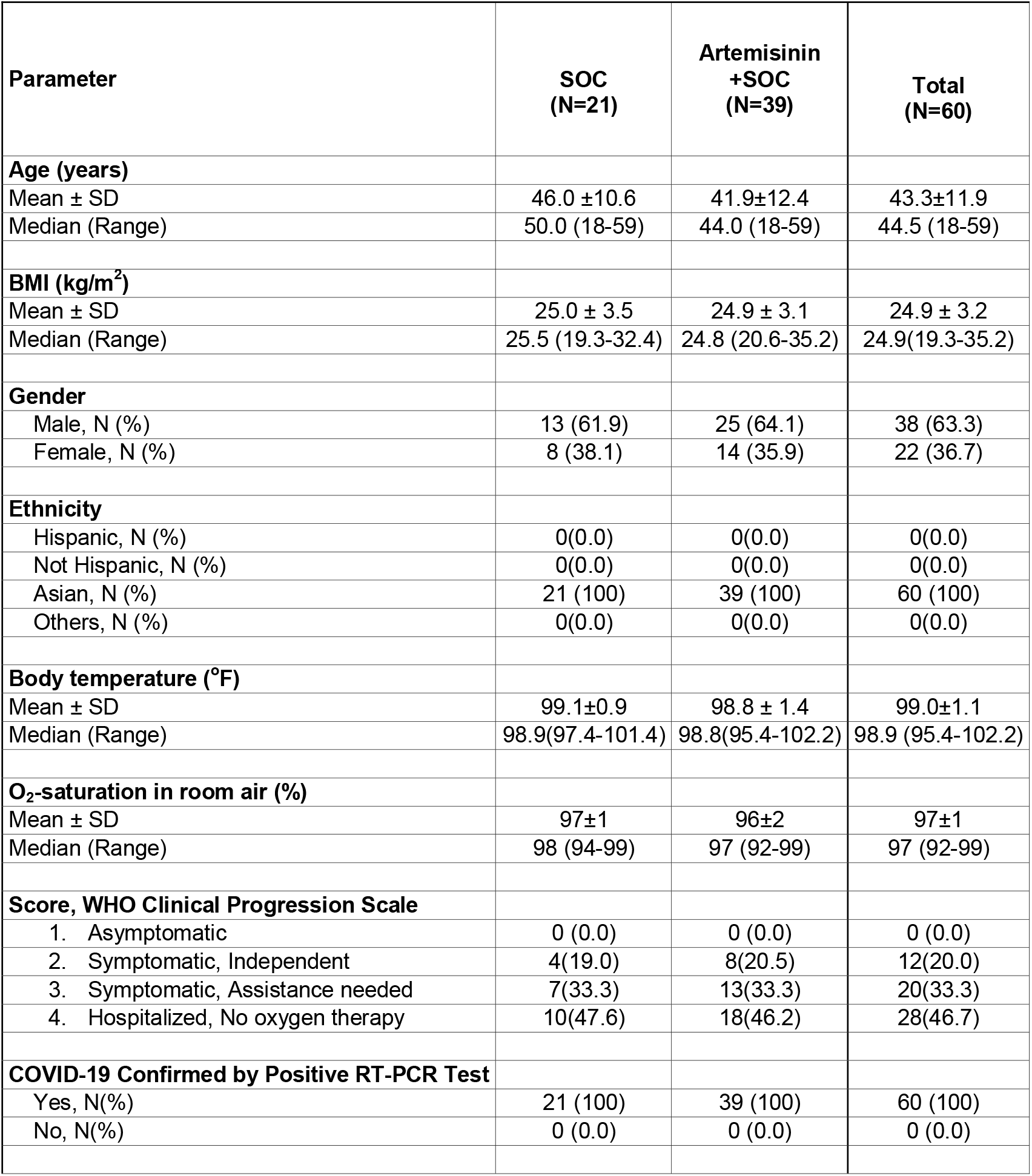
Patient Demographics and Baseline Characteristics.

| Parameter | SOC<br>(N=21) | Artemisinin<br>+SOC<br>(N=39) | Total<br>(N=60) |
| --- | --- | --- | --- |
| **Age (years)** |  |  |  |
| Mean ± SD | 46.0 ± 10.6 | 41.9 ± 12.4 | 43.3 ± 11.9 |
| Median (Range) | 50.0 (18-59) | 44.0 (18-59) | 44.5 (18-59) |
| **BMI (kg/m<sup>2</sup>)** |  |  |  |
| Mean ± SD | 25.0 ± 3.5 | 24.9 ± 3.1 | 24.9 ± 3.2 |
| Median (Range) | 25.5 (19.3-32.4) | 24.8 (20.6-35.2) | 24.9 (19.3-35.2) |
| **Gender** |  |  |  |
| Male, N (%) | 13 (61.9) | 25 (64.1) | 38 (63.3) |
| Female, N (%) | 8 (38.1) | 14 (35.9) | 22 (36.7) |
| **Ethnicity** |  |  |  |
| Hispanic, N (%) | 0(0.0) | 0(0.0) | 0(0.0) |
| Not Hispanic, N (%) | 0(0.0) | 0(0.0) | 0(0.0) |
| Asian, N (%) | 21 (100) | 39 (100) | 60 (100) |
| Others, N (%) | 0(0.0) | 0(0.0) | 0(0.0) |
| **Body temperature (°F)** |  |  |  |
| Mean ± SD | 99.1 ± 0.9 | 98.8 ± 1.4 | 99.0 ± 1.1 |
| Median (Range) | 98.9 (97.4-101.4) | 98.8 (95.4-102.2) | 98.9 (95.4-102.2) |
| **O<sub>2</sub>-saturation in room air (%)** |  |  |  |
| Mean ± SD | 97 ± 1 | 96 ± 2 | 97 ± 1 |
| Median (Range) | 98 (94-99) | 97 (92-99) | 97 (92-99) |
| **Score, WHO Clinical Progression Scale** |  |  |  |
| 1. Asymptomatic | 0 (0.0) | 0 (0.0) | 0 (0.0) |
| 2. Symptomatic, Independent | 4 (19.0) | 8 (20.5) | 12 (20.0) |
| 3. Symptomatic, Assistance needed | 7 (33.3) | 13 (33.3) | 20 (33.3) |
| 4. Hospitalized, No oxygen therapy | 10 (47.6) | 18 (46.2) | 28 (46.7) |
| **COVID-19 Confirmed by Positive RT-PCR Test** |  |  |  |
| Yes, N(%) | 21 (100) | 39 (100) | 60 (100) |
| No, N(%) | 0 (0.0) | 0 (0.0) | 0 (0.0) |

### Safety

As of the data cut-off date, safety data were available for all 60 participants, including 39 patients who received ARTIVeda. All AEs encountered in all 60 patients are shown in **Table 2** according to their SOC, severity, and relatedness to the assigned treatments. There were no incidences of drug-related deaths or discontinuation of therapy. No treatment-related ≥Grade 3 AEs were observed in any of the patients. The only ARTIVeda-related AEs were a single occurrence of transient mild rash and mild vertigo (**Table 2**). These AEs did not require additional treatment, and they did not cause discontinuation of the dosing or study. All AEs considered possibly or probably related to ARTIVeda have recovered/resolved with no sequelae. No laboratory abnormalities were observed in the SOC plus ARTIVeda group to suggest drug-related systemic or organ-specific toxicity. There were no clinically meaningful changes in laboratory values identified in observed values from baseline compared with the SOC alone group (**Table 3**). In particular, laboratory tests performed on day 28 showed: (*a*) no evidence of kidney toxicity, as documented by their normal serum BUN and creatinine levels; (b) no evidence of liver toxicity, as documented by their normal ALT, AST, and total bilirubin levels, and (*b*) no evidence of hematological toxicity, as documented by their normal and near-baseline RBC counts, hemoglobin, platelet counts, and WBC counts with normal percentages of neutrophils and lymphocytes.

**Table 2.**
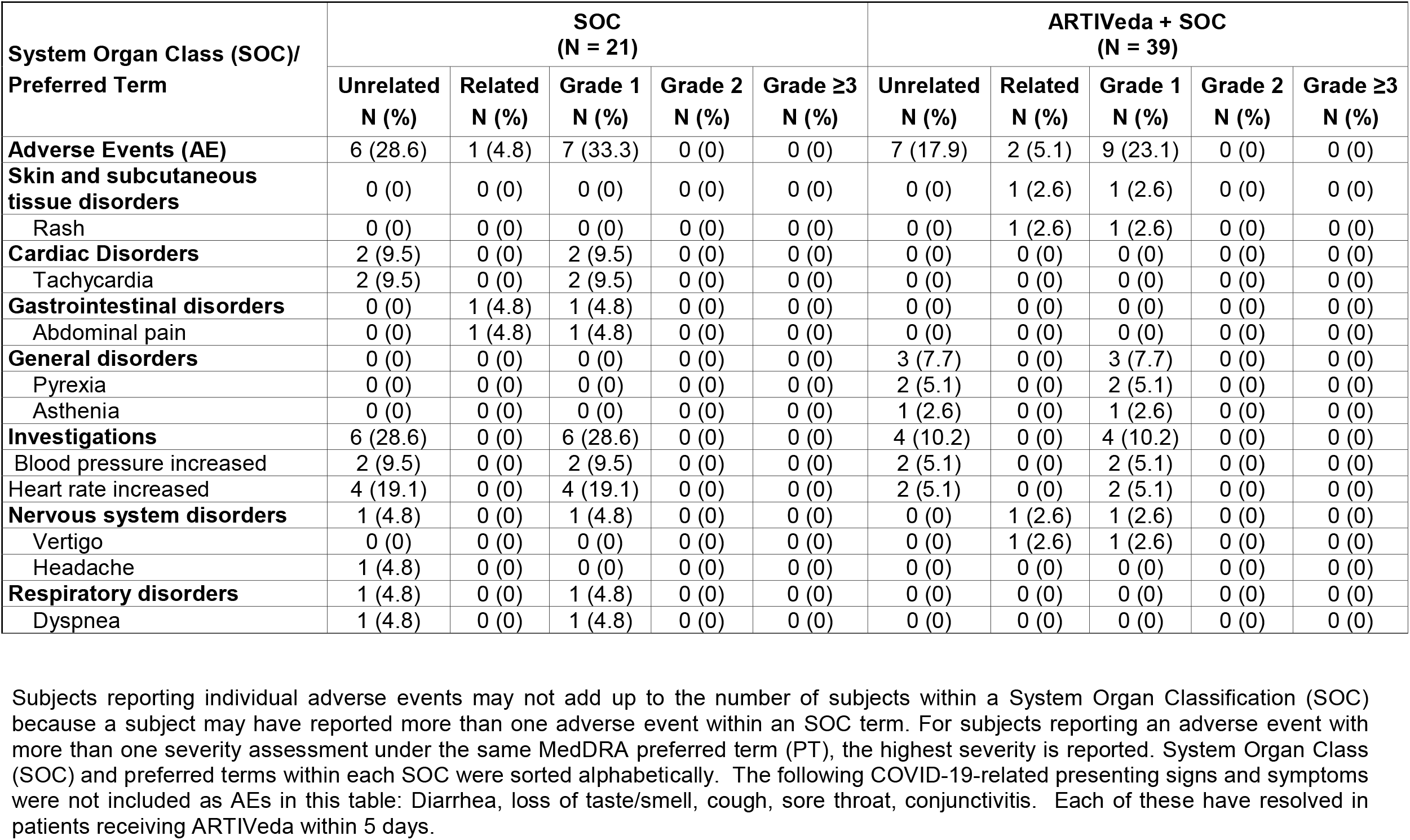
Adverse Events Reported for All Patients Treated with ARTIVeda on the ARTI-19 Study.

| System Organ Class (SOC)/<br>Preferred Term | SOC<br>(N = 21) |  |  |  |  | ARTIVeda + SOC<br>(N = 39) |  |  |  |  |
| --- | --- | --- | --- | --- | --- | --- | --- | --- | --- | --- |
|  | Unrelated<br>N (%) | Related<br>N (%) | Grade 1<br>N (%) | Grade 2<br>N (%) | Grade ≥3<br>N (%) | Unrelated<br>N (%) | Related<br>N (%) | Grade 1<br>N (%) | Grade 2<br>N (%) | Grade ≥3<br>N (%) |
| **Adverse Events (AE)** | 6 (28.6) | 1 (4.8) | 7 (33.3) | 0 (0) | 0 (0) | 7 (17.9) | 2 (5.1) | 9 (23.1) | 0 (0) | 0 (0) |
| **Skin and subcutaneous**<br>**tissue disorders** | 0 (0) | 0 (0) | 0 (0) | 0 (0) | 0 (0) | 0 (0) | 1 (2.6) | 1 (2.6) | 0 (0) | 0 (0) |
| Rash | 0 (0) | 0 (0) | 0 (0) | 0 (0) | 0 (0) | 0 (0) | 1 (2.6) | 1 (2.6) | 0 (0) | 0 (0) |
| **Cardiac Disorders** | 2 (9.5) | 0 (0) | 2 (9.5) | 0 (0) | 0 (0) | 0 (0) | 0 (0) | 0 (0) | 0 (0) | 0 (0) |
| Tachycardia | 2 (9.5) | 0 (0) | 2 (9.5) | 0 (0) | 0 (0) | 0 (0) | 0 (0) | 0 (0) | 0 (0) | 0 (0) |
| **Gastrointestinal disorders** | 0 (0) | 1 (4.8) | 1 (4.8) | 0 (0) | 0 (0) | 0 (0) | 0 (0) | 0 (0) | 0 (0) | 0 (0) |
| Abdominal pain | 0 (0) | 1 (4.8) | 1 (4.8) | 0 (0) | 0 (0) | 0 (0) | 0 (0) | 0 (0) | 0 (0) | 0 (0) |
| **General disorders** | 0 (0) | 0 (0) | 0 (0) | 0 (0) | 0 (0) | 3 (7.7) | 0 (0) | 3 (7.7) | 0 (0) | 0 (0) |
| Pyrexia | 0 (0) | 0 (0) | 0 (0) | 0 (0) | 0 (0) | 2 (5.1) | 0 (0) | 2 (5.1) | 0 (0) | 0 (0) |
| Asthenia | 0 (0) | 0 (0) | 0 (0) | 0 (0) | 0 (0) | 1 (2.6) | 0 (0) | 1 (2.6) | 0 (0) | 0 (0) |
| **Investigations** | 6 (28.6) | 0 (0) | 6 (28.6) | 0 (0) | 0 (0) | 4 (10.2) | 0 (0) | 4 (10.2) | 0 (0) | 0 (0) |
| Blood pressure increased | 2 (9.5) | 0 (0) | 2 (9.5) | 0 (0) | 0 (0) | 2 (5.1) | 0 (0) | 2 (5.1) | 0 (0) | 0 (0) |
| Heart rate increased | 4 (19.1) | 0 (0) | 4 (19.1) | 0 (0) | 0 (0) | 2 (5.1) | 0 (0) | 2 (5.1) | 0 (0) | 0 (0) |
| **Nervous system disorders** | 1 (4.8) | 0 (0) | 1 (4.8) | 0 (0) | 0 (0) | 0 (0) | 1 (2.6) | 1 (2.6) | 0 (0) | 0 (0) |
| Vertigo | 0 (0) | 0 (0) | 0 (0) | 0 (0) | 0 (0) | 0 (0) | 1 (2.6) | 1 (2.6) | 0 (0) | 0 (0) |
| Headache | 1 (4.8) | 0 (0) | 0 (0) | 0 (0) | 0 (0) | 0 (0) | 0 (0) | 0 (0) | 0 (0) | 0 (0) |
| **Respiratory disorders** | 1 (4.8) | 0 (0) | 1 (4.8) | 0 (0) | 0 (0) | 0 (0) | 0 (0) | 0 (0) | 0 (0) | 0 (0) |
| Dyspnea | 1 (4.8) | 0 (0) | 1 (4.8) | 0 (0) | 0 (0) | 0 (0) | 0 (0) | 0 (0) | 0 (0) | 0 (0) |

Subjects reporting individual adverse events may not add up to the number of subjects within a System Organ Classification (SOC) because a subject may have reported more than one adverse event within an SOC term. For subjects reporting an adverse event with more than one severity assessment under the same MedDRA preferred term (PT), the highest severity is reported. System Organ Class (SOC) and preferred terms within each SOC were sorted alphabetically. The following COVID-19-related presenting signs and symptoms were not included as AEs in this table: Diarrhea, loss of taste/smell, cough, sore throat, conjunctivitis. Each of these have resolved in patients receiving ARTIVeda within 5 days.

**Table 3.**
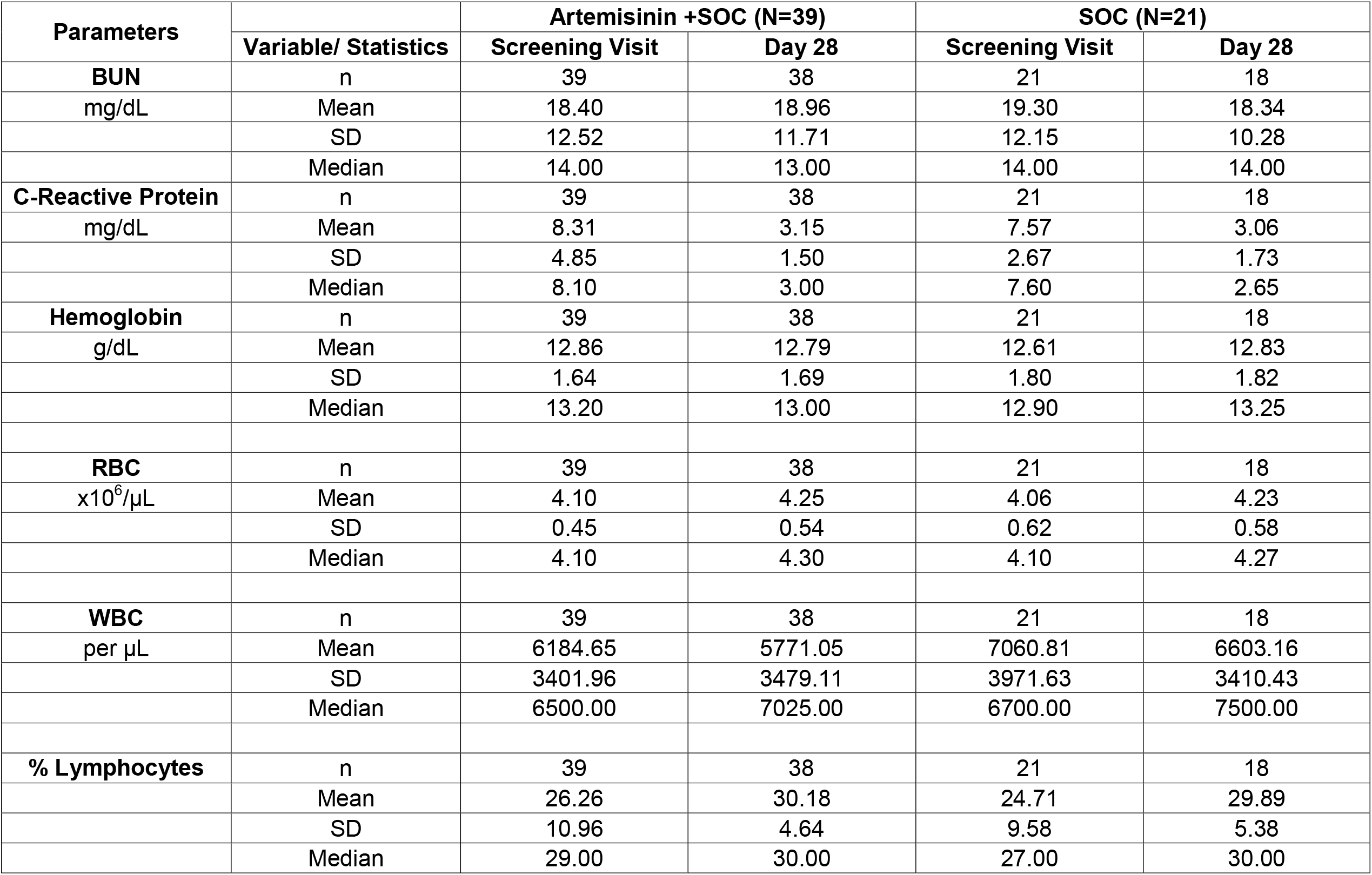
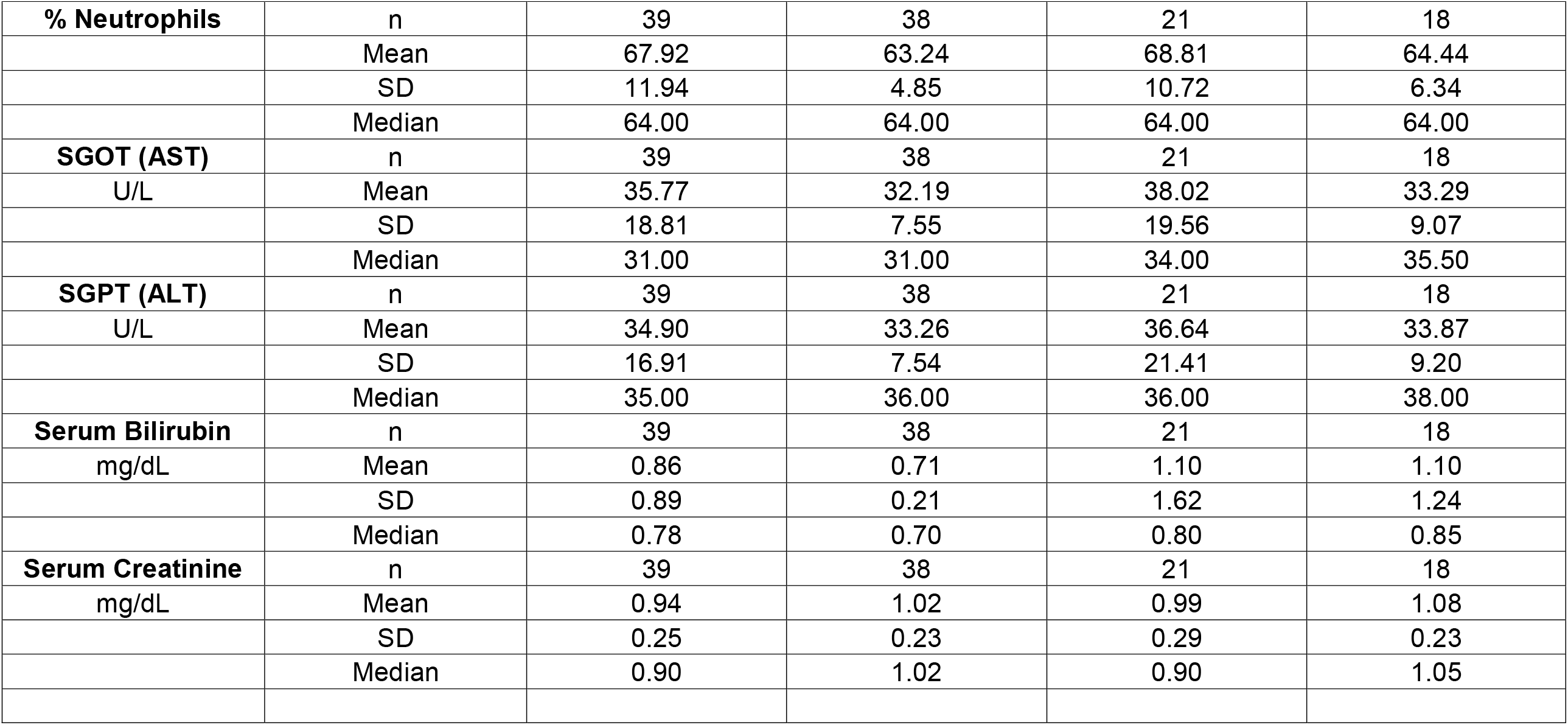
Safety Laboratory Test Results.

### Efficacy

More patients recovered faster when ARTIVeda was used as adjunct therapy alongside SOC. This was observed across all sites including Remdesivir-based SOC at site 201, Dexamethasone/Heparin-based SOC at site 202, and Ivermectin-based SOC at site 203. While all patients were symptomatic at baseline (WHO score = 2-4), 31 of 39 (79.5%) of patients treated with ARTIVeda plus SOC became asymptomatic (WHO score = 1) by the end of the 5-day therapy, including 10 of 10 patients with severe dry cough 7 of 7 patients with severe fever. By comparison, 12 of 21 control patients (57.1%) treated with SOC alone became asymptomatic on day 5 (P=0.028, Fisher’s exact test) (**Table 4**). These results indicate that ARTIVeda when added to the SOC accelerates the recovery of patients with mild-moderate COVID-19. This clinical benefit was particularly evident when the treatment outcomes of hospitalized COVID-19 patients (WHO score = 4) treated with SOC alone versus SOC plus ARTIVeda were compared. The median time to becoming asymptomatic was only 5 days for the SOC plus ARTIVeda group (N=18) but 14 days for the SOC alone group (N=10) (P=0.004, Log-rank test) (**Figure 1**). While 11 of 18 (61.1%) hospitalized patients treated with SOC plus ARTIVeda became asymptomatic by day 5, only 2 of 10 (20%) of hospitalized patients treated with SOC alone did (P=0.04, Fisher’s exact test).

**Table 4.** ARTIVeda Accelerates Recovery of Patients with Mild-Moderate COVID-19.

| Treatment | Score* | Baseline | EOT<br>Day 5 | Day 14 | Day 28 |
| --- | --- | --- | --- | --- | --- |
| **Experimental arm,**<br>**ARTIVeda + SOC**<br>**N (%)** | 1 | 0 (0) | **31 (79.5)** | 39 (100) | 39 (100) |
|  | 2 | 8 (20.5) | 4 (10.3) | 0 (0) | 0 (0) |
|  | 3 | 13 (33.3) | 4 (10.3) | 0 (0) | 0 (0) |
|  | 4 | 18 (46.2) | 0 (0) | 0 (0) | 0 (0) |
| **Control arm, SOC**<br>**N (%)** | 1 | 0 (0) | **12 (57.1)** | 21 (100) | 21 (100) |
|  | 2 | 4 (19.0) | 8 (38.1) | 0 (0) | 0 (0) |
|  | 3 | 7 (33.3) | 1 (4.8) | 0 (0.0) | 0 (0) |
|  | 4 | 10 (47.6) | 0 (0) | 0 (0) | 0 (0) |
| P-value; Score 1,<br>Experimental arm vs. Control<br>arm, Fisher's exact test | - | - | **P=0.028** | - | - |

\*Score according to the 10-point WHO clinical progression scale. 1 = asymptomatic; 2=symptomatic, independent; 3=symptomatic requiring assistance; 4=hospitalized, not requiring oxygen therapy.

**Figure 1.**
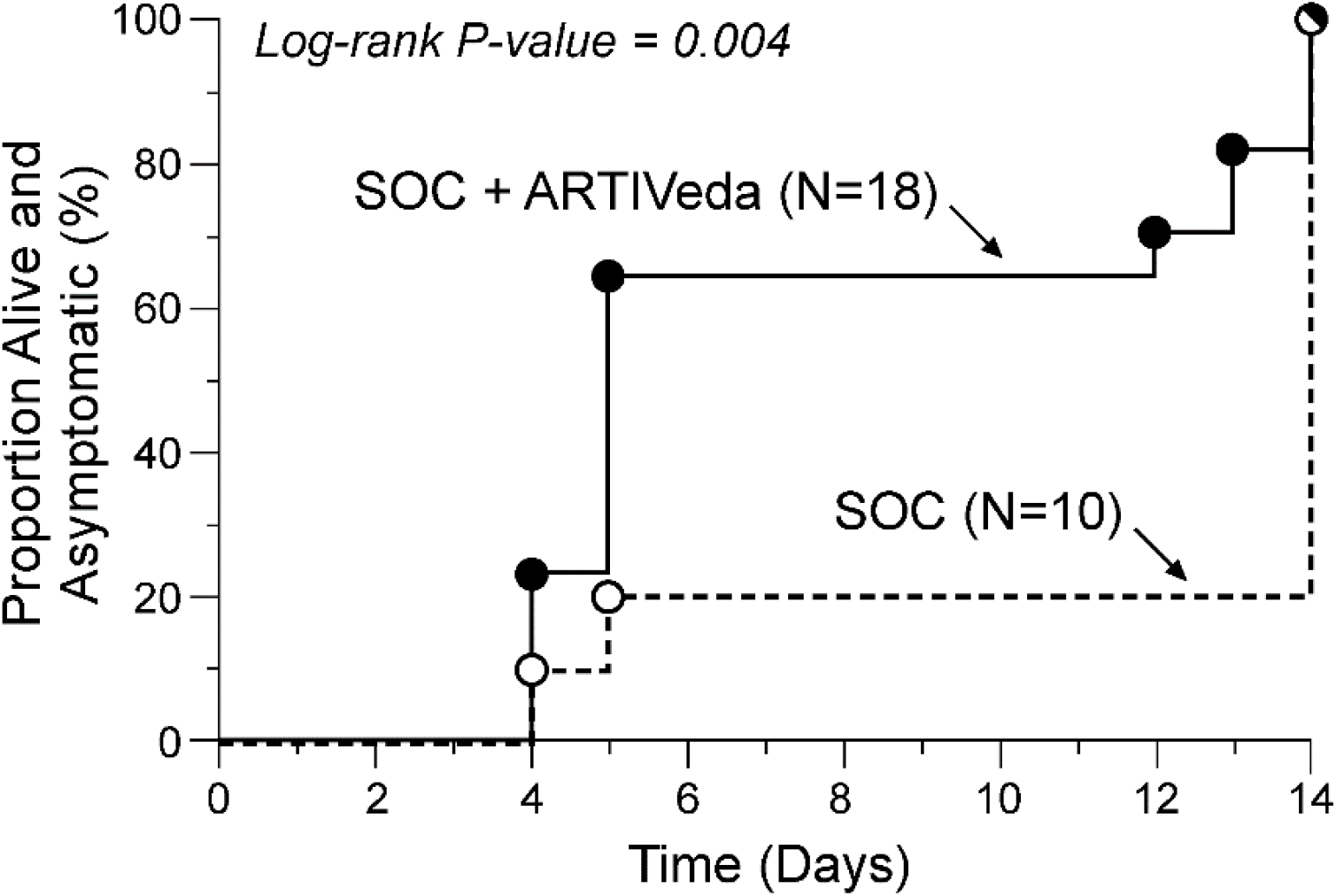
ARTIVeda Accelerates Recovery of Patients with Moderate COVID-19. Depicted are the recovery curves showing the time to becoming asymptomatic for the SOC alone and SOC plus ARTIVeda groups along with the log-rank P-value for the comparison.

## DISCUSSION

*Artemisia* species contain bioactive substances with pleiotropic biological effects (10, 12). For example, *Artemisia annua* contains anti-inflammatory sesquiterpenoids, including Artemisinin (viz.: Artesunate). Artemisinin and its antimalarial properties were discovered by the Chinese scientist Tu Youyu, who became one of the laureates of the 2015 Nobel Prize in Physiology or Medicine for this discovery (12). Artemisinin and Artemisinin derivatives are generally well-tolerated, especially when used for a short treatment course (10, 12, 18-24). Except for the rare occurrence of hepatotoxicity and mild-moderate headache, nausea, vomiting, fatigue, and anorexia, Artemisinin was found to be clinically safe in healthy volunteers as well as malaria patients (10, 12, 18-24). In the present study, ARTIVeda was administered daily for 5 days as an adjunct to the Remdesivir-based SOC at Site 201, Dexamethasone/Heparin-based SOC at Site 202, or Ivermectin-based SOC at Site 203. No treatment-related ≥Grade 3 AEs were observed in any of the patients. The only ARTIVeda-related AEs were mild rash and mild hypertension, and these AEs did not require additional treatment, and they did not cause discontinuation of the dosing or study. All AEs considered possibly or probably related to ARTIVeda have recovered/resolved with no sequelae. No laboratory abnormalities were observed in the SOC plus ARTIVeda group to suggest drug-related systemic or organ-specific toxicity. Most importantly, no hepatotoxicity was observed in any of the 39 patients treated with ARTIVeda.

Artemisinin and some of its chemical derivatives exhibit broad-spectrum antiviral activity against pathogenic human viruses, including SARS-CoV-2 (11, 12). Artemisinin has also been shown to inhibit the expression of TGF-β mRNA and protein both in vitro and in vivo (25-27). Artemisinin treatments mitigated the contribution of upregulated TGF-β expression to the disease pathophysiology in rodent models of diabetic nephropathy as well as lupus nephritis (26, 27). High-risk COVID-19 patients have a higher probability of developing a potentially life-threatening multi-system inflammation caused by a cytokine release syndrome (CRS) (4-9). TGF-β is one of the important pro-inflammatory cytokines that contribute to the inflammatory injury of lungs in COVID-19 patients during the CRS (28-31). Notably, infection with SARS-CoV increases the expression of TGF-β and potentiates the TGF-β-regulated MAPK-mediated inflammatory signals (32-35). The upregulation of TGF-β genes in COVID-19 patients has also been documented (31). TGF-β also worsens the severity of pulmonary edema and acute respiratory distress syndrome (ARDS) that develops after CRS by increasing capillary permeability and impairing alveolar fluid transportation in the lungs (36-40). In addition, neutrophil extracellular traps (NETs) that are formed via activation of the complement system and production of complement cleavage products during sepsis trigger TGF-β release from platelets and thereby contribute to persistence of systemic inflammation and serious coagulopathy (41-45). Due to the pivotal role of TGF-β in the pathophysiology of lung fibrosis that develops after inflammatory injury to the lungs (46-49), the use of a TGF-β inhibitor like Artemisinin has the clinical potential to prevent pulmonary fibrosis in COVID-19 patients. Our results presented herein provide clinical proof of concept that targeting the TGF-β pathway with ARTIVeda may contribute to a faster recovery of patients with mild-moderate COVID-19 when administered early in the course of their disease.

Artemisinin and some of its derivatives are being evaluated as COVID-19 therapeutics (12). ArtemiC is a medical spray containing Artemisinin, curcumin, Frankincense resin from the Boswellia sacra tree, and Vitamin C. In the controlled Phase II trial NCT04382040, patients with COVID-19 received ArtemiC spray in addition to standard care and preliminary data suggested that ArtemiC may be more active than placebo in contributing to the improvement of the patients’ condition (50). Likewise, the efficacy signal for the Artemisinin derivative Artesunate during a recently completed prospective, controlled clinical COVID-19 study was promising. In Artesunate treatment group, time to significant improvement of the symptoms, time to conversion to the negativity of SARS-CoV-2 tests, and length of hospital stay was shorter than in the control group (51). Additional data sets are expected from several ongoing active clinical trials designed to evaluate the efficacy of Artemisinin and its derivatives in COVID-19 patients, including NCT04387240 Phase 2 trial testing the efficacy of Artesunate and Artemisinin will be examined in patients with mild COVID-19, NCT04502342 trial testing the efficacy of cospherunate, a combination of Artesunate and amodiaquine, NCT04475107 Phase 2/3 trial testing the efficacy of pyramax, a combination of pyronaridine and Artesunate, IRCT20200607047682N1 Phase 2/3 trial testing the efficacy of Anval S (an oral syrup containing Artemisinin) plus Azithromycin, NCT04553705 trial testing the efficacy of Artemisinin combined with Omega-3 supplementation and thymoquinone, NCT04374019 Phase 2 trial testing the efficacy of Artemisia annua as well as Artesunate in high-risk COVID-19 patients.

In summary, Artemisinin-containing drugs, such as ARTIVeda, have clinical impact potential in the treatment of COVID-19 because it can prevent the progression of the disease and accelerate the recovery of patients before they develop potentially life-threatening complications. This dual-function COVID-19 drug candidate is hoped to mitigate the TGF-b mediated inflammatory injury associated with the cytokine storm and viral sepsis in critically ill COVID-19 patients. Our study provides the rationale for further clinical development of ARTIVeda as a COVID-19 drug candidate. Based on the encouraging activity signal in mild-moderate COVID-19 patients, we are planning to start the clinical testing of ARTIVeda in severe to critically ill COVID-19 patients as well.

## Data Availability

Data available upon request.

## Author contributions

Each author has made significant and substantive contributions to the study, reviewed and revised the manuscript, provided final approval for submission of the final version. No medical writer was involved. V.T and F.M.U conceived the study, designed the evaluations reported in this paper, directed the data compilation and analysis, analyzed the data, and prepared the initial draft of the manuscript. Each author had access to the source data used in the analyses.

